# Utilizing LLMs for Enhanced Argumentation and Extraction of Causal Knowledge from Scientific Literature

**DOI:** 10.1101/2024.03.20.24304652

**Authors:** Shuang Wang, Wenjie Chen, Yang Zhang, Ting Chen, Jian Du

## Abstract

Current semantic extraction tools have limited performance in identifying causal relations, neglecting variations in argument quality, especially persuasive strength across different sentences. The present study proposes a five-element based (evidence cogency, concept, relation stance, claim-context relevance, conditional information) causal knowledge mining framework and automatically implements it using large language models (LLMs) to improve the understanding of disease causal mechanisms. As a result, regarding cogency evaluation, the accuracy (0.84) of the fine-tuned Llama2-7b largely exceeds the accuracy of GPT-3.5 turbo with few-shot. Regarding causal extraction, by combining PubTator and ChatGLM, the entity first-relation later extraction (recall, 0.85) outperforms the relation first-entity later means (recall, 0.76), performing great in three outer validation sets (a gestational diabetes-relevant dataset and two general biomedical datasets), aligning entities for further causal graph construction. LLMs-enabled scientific causality mining is promising in delineating the causal argument structure and understanding the underlying mechanisms of a given exposure-outcome pair.

## Introduction

The scientific literature is rich with information across diverse fields concerning potential disease mechanisms, e.g., causal knowledge surrounding diseases. However, identifying and prioritizing more promising and reliable mechanisms for further analytical evaluation is a great challenge for medical researchers, with the increasing growth of biomedical literature and related datasets (1). Constructing knowledge graphs by extracting concepts and their relations from scientific texts effectively solves this challenge. For example, the Semantic MEDLINE Database (SemMedDB) is a representative repository of subject-predicate-object triples extracted from the entire set of PubMed titles and abstracts (2). SemMedDB has been utilized by manually selecting *casual/mechanistic relations such* as CAUSES, PREVENTS, and DISRUPTS to identify intermediates (3-5) and common causes, i.e., confounders (6, 7) between the investigated exposure-and-outcome variables. While widely used for constructing biomedical causal knowledge graphs, it demonstrated limited performance with precision of 0.69, recall of 0.42, and an F1 score of 0.52 in a relaxed evaluation (8). Even using a relation classification approach to eventually increase the size and the utility of SemMedDB, the best model yields a precision of 0.62, and a recall of 0.81(9). This would highly influence the quality of its downstream task. For instance, since the concepts extracted are overly general, it can result in many apparent contradictions that are not truly contradictory (10). Additionally, existing natural language processing (NLP) techniques for biomedical entity and relations extraction frequently neglect the quality of arguments, especially the varying degrees of strength across different sentences where claims are expressed. While seven factuality values (fact, probable, possible, doubtful, counterfact, uncommitted, and conditional) were annotated for the extracted triples, representing the real-world nature of biomedical knowledge as hypotheses, speculations, or opinions rather than explicit facts (11), this only captures one facet of argument quality, i.e., the uncertainty in scientific discourse.

In this study, we introduce the concept of argument quality to enhance the understanding of causality for disease mechanisms mining and evidence-based clinical decision-making. Argumentation plays a fundamental role in complex areas like healthcare, where it directly affects decision-making related to human lives. Argument mining, a method in computational linguistics, has been a significant technique in evidence-based decision-making to use the best evidence to improve the quality of medical research as well as clinical practice (12, 13). Recently, an impressive development in argument mining is Project Debater (14, 15). The training corpus curated by IBM Project Debater is organized under a simple claim-evidence structure, allowing for the automatic detection of claims and evidence in the context of Wikipedia’s controversial topics (16). Project Debater focuses on social issues, and the basic structure of a claim is “Topic-Action” or “Topic-View”. For example, a policy motion like “Preschool should be subsidized”, where the topic is “Preschool”, and the action is “be subsidized”; a legitimate analysis motion like “AI brings more harm than good”, where the topic is “AI”, and the view is “brings more harm than good”, or the opposite stance. In this study, we mainly focus on scientific claims in biomedical fields instead of social issues. Compared with societal claims, scientific claims mainly represented a Cause-Effect structure, such as *Cancer_COMPLICATES_Covid-19*, i.e., “Cancer patients are susceptible to COVID-19”, and *Hydroxychloroquine_NEG TREATS_Covid-19*, i.e., “hydroxychloroquine should not be used for patients with COVID-19 of any severity”. Here we consider an extracted triple as a claim, its multiple sources of supporting sentences as the evidence set. While this is a simplified model, it captures the essence of argument mining.

Here, we link causal mining to the thought of argument mining, through which a causal statement can be considered as a common structured argument consisting of a premise (evidence/cause) and a conclusion (effect). Thus, the essence of causality extraction is to identify conclusive semantic relations in an argument. However, not all arguments hold equal strength or persuasion (17). Various dimensions, such as cogency, reasonableness, and effectiveness, have been proposed to assess argumentative quality. Moreover, evaluating argument quality presents a formidable challenge. For instance, determining logical cogency requires analyzing the acceptability, relevance, and collective sufficiency of premises, demanding close reading and logical reasoning.

Large Language Models (LLMs) have demonstrated substantial potential across diverse applications in the medical field. We agree with such viewpoint that, it is more appropriate to employ LLMs as scientific reasoning engines, rather than knowledge bases since they often fall short when it comes to the factual accuracy of the generated scientific knowledge (18). We assume LLMs would perform well in such a complex task of causal argument mining, given the multitude of logical reasoning processes involved, including assessing the varying strength of evidence across different sentences and discerning strong conclusive correlation in extracted triplets within sentences, thus minimizing information loss. Thus, the present study aims to utilize LLMs to design a pipeline framework to automatically extract causal triples for the facilitated capture and understanding of causal knowledge, e.g., disease mechanisms mined from the scientific literature.

## Method

First, after multiple rounds of annotation and adjustment, we proposed a causal argument mining framework containing five key elements, evidence cogency, concept, relation stance, claim-context relevance, and conditional information. Second, based on the framework, we utilize LLMs to extract causal triples automatically. Third, we evaluate the models’ performance using three outer validation sets, including a dataset associated with gestational diabetes (GDM) and two general biomedical datasets. The project workflow is shown below (Figure 1).

**Figure 1.**
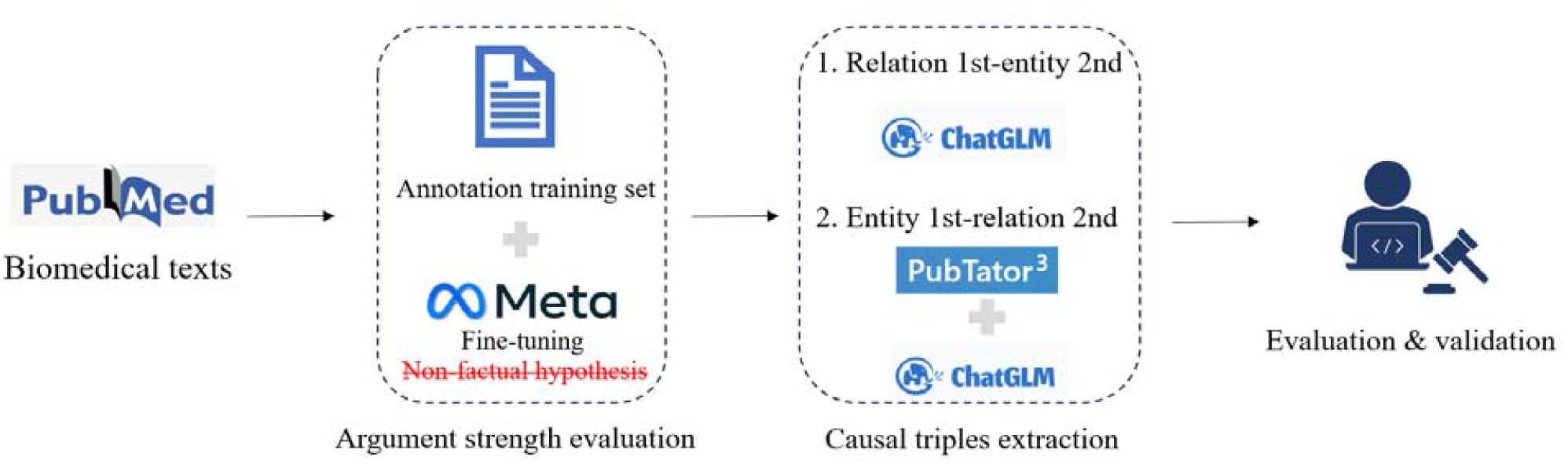
The research workflow of LLMs-enabled causal extraction

### 1. The framework of causal argument mining

Inspired by the classic argument mining methods, to automatically target causal argument relations from texts, we design the framework into three major steps (Figure 2). First, identify causal sentences from texts. Second, identify the causal argument component in which we focus on conclusive components. Third, identify other conditional information.

**Figure 2.**
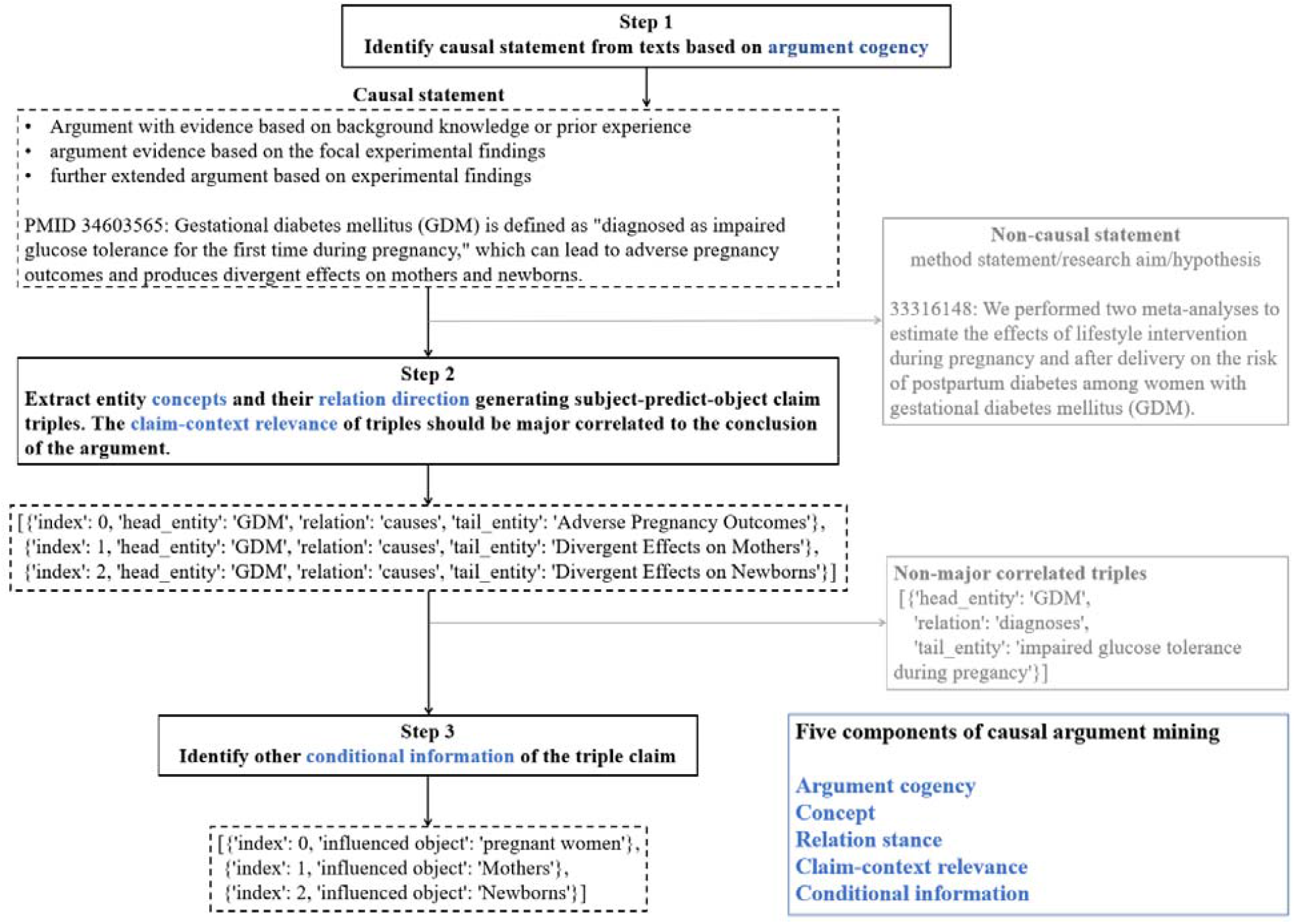
The framework of five elements of causal argument mining

#### 1) Measure argument cogency

To conduct causal argument mining from free texts, the initial step is to identify causal sentences. As it is difficult to define a causal sentence, we conduct causal mining from the perspective of argument mining. In general, a causal statement can be considered as a simple structured evidence-based argument consisting of a premise (evidence/cause) and a conclusion (effect). Thus, based on the extent of evidence cogency (the logical strength of evidence to support the claim), we classify argument cogency into four categories, 1) argument lack of supporting information (i.e., research hypothesis, research design), 2) argument with evidence based on background knowledge or prior experience, 3) argument evidence based on the focal experimental findings, and 4) further extended argument based on experimental findings. To identify causal arguments, we remove category 1 which are weak arguments without clear opinions or those without supporting evidence.

#### 2) Identify the causal argument component

After identifying a casual statement, we then extract key causal relations involved by identifying conclusive subject-predicate-object triples. We first examine whether a triple claim is accurately extracted from the source causal evidence sentence. Then we measure the claim-evidence correlation to classify a triple claim as a premise or a conclusion and keep only conclusive triples as causal claim triples.

##### Concept and relation stance of a claim triple

A causal sentence may involve many flat triples. Each extracted triple consists of head and tail concepts, as well as the predicate direction. Regarding predicate, we manually design a list of causal-related predicates (N = 10, e.g., *CAUSES* and *PREDISPOSES*) as well as their causal direction, which are inhibitory, excitatory, and neutral. A neutral direction may be extracted from the conditional statement rather than an assertion.

##### Claim-context relevance

Relevance degree measures whether 1) a triple is connected to the main assertion of the original sentence as a conclusion (major relevance) or 2) a triple involved in the sentence as an implied premise (non-major relevance).

#### 3) Identify conditional information

A notable concern with the subject-predict-object triple is its flat structure. It only represents the smallest unit of factual information without detailed conditional information and hierarchical structures, especially the complex phenotypes and influential populations of diseases. For example, as we choose GDM-relevant research sentences as a validation set, identifying subsequent outcomes for the influenced object, especially the maternal or fetus, is an essential problem. GDM can not only lead to obesity in the mother but also the child born. Extracting and representing the conditional information of triples is a highly complex task, given that conditions can arise from various sources. Presently, there is ongoing studies focused on the extraction of such conditional information in biomedical literature (19) and clinical trials (20). To examine the potential of LLMs in supplementing conditional information, here, we specify the conditional information as the influenced object of the given triple and utilize LLMs to extract such information as a supplement for further refined data integration.

### 2. Automatic extraction using LLMs

#### 1) The automatic identification of argument cogency

Evidence cogency is essentially a complex task for models. According to the initial interaction with GPT-4 (21), we find that it is hard to distinguish background knowledge from research experimental findings completely. Even optimizing a detailed prompt with explanations and differential definitions and few-shot (9-shot) using GPT-3.5-turbo (21), as the prediction data size increases, models often mix methodology statements with new findings, which suggests the need for an advanced training technique. Thus, we decided to fine-tune a lightweight model for the cogency evaluation task. We annotated 576 sentences as the training set and then fine-tuned Llama2-7b (22) for the cogency task. For performance evaluation, we use few-shot GPT-3.5-turbo as a baseline model, based on a separate annotation test set (N = 100), we compared the accuracy and macro F1 scores of fine-tuned Llama2-7b and the baseline model.

#### 2) Extraction strategies

We then conduct the automatic causal triple extraction based on the designed casual argument mining framework. With ChatGLM (23) as the extraction model, we compared two extraction strategies: 1) Relation first – entity later (relation-entity), and 2) Entity first – relation later (entity-relation) (Figure 3). As for strategy 1, in a given casual statement, we have ChatGLM first identify whether any casual relations exist in the sentence and then identify each relation’s head and tail entities. Strategy 2 reverses the two steps. Firstly, PubTator (24), a named entity recognition (NER) tool that can identify standard concepts of Gene, Disease, Chemical, Species, CellLine, and Variant for biomedical texts, is used to extract all biomedical entities in the sentence, and then ChatGLM identifies existed casual relations in any two entities generating casual triples involved. For extraction evaluation, as there are no gold standard datasets of the casual triples’ extraction, we manually made a GDM-related test set for performance evaluation and included two NLM datasets as independent outer validation datasets. In specific, we manually annotated casual triples of 222 GDM-related sentences as the test. Also, we filtered and included merely casual triple-casual statements from a human-annotated NLM gold standard triple-sentence dataset (N = 142) as the outer validation set. Another SemRep automatic triple-sentence extraction dataset (N = 1,635) is also involved as an outer validation set for comparison and assessment.

**Figure 3.**
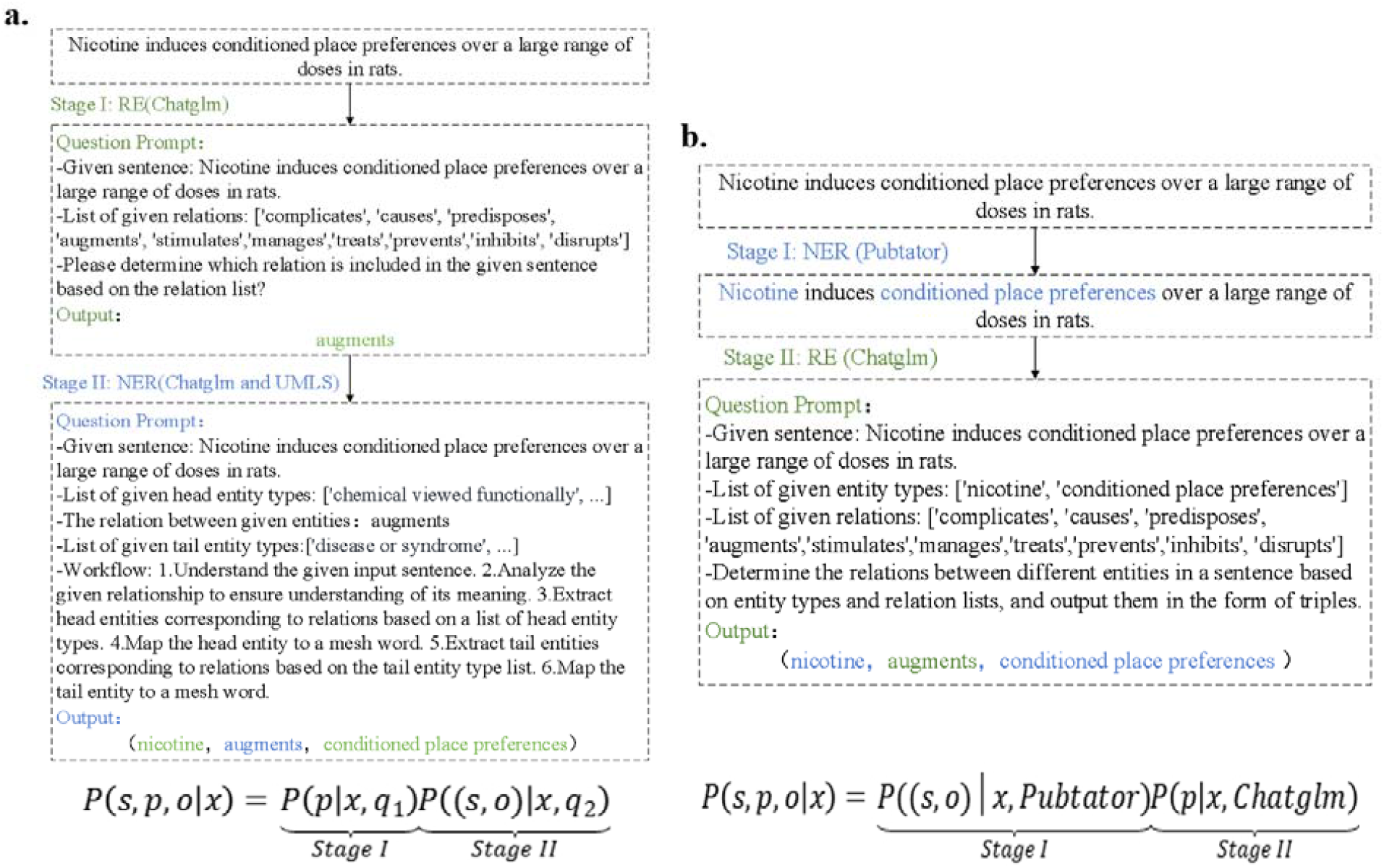
LLMs-based causal extraction workflow. **a**. The causal extraction workflow using the method of relation 1^st^-entity 2^nd^ (ChatGLM). **b**. The causal extraction workflow using the method of entity 1^st^ (PubTator)-relation 2^nd^ (ChatGLM).

## Results

First of all, in measuring argument cogency, with 576 GDM relevant annotated sentences as a training set, the fine-tuned Llama2-7b exhibits great performance scores with an accuracy of 0.84, highly superior to few-shot GPT-3.5 with an accuracy of 0.44 (Table 1-A). The accurate identification of causal statements guarantees the proceeding downstream tasks of causal mining. Second, in automatically extracting causal triples, both extraction approaches show relatively high scores in either the GDM test dataset or the NLM gold standard dataset. This reflects the robust performance of two extraction methods in the GDM field and its generalizability in general biomedical texts. In particular, the overall performance in all test sets reveals that the entity-relation means exceed the relation-entity method. In addition, when using the NLM SemRep extraction dataset as true values for testing, the relatively lower recall (65.32%, 70.64%) and precision (66.67%, 71.21%) scores than that of the NLM manual-curated gold standard dataset suggest that our ChatGLM-based extraction approach precedes SemRep.

**Table 1.** Performance evaluation results of LLM-based causal extraction.

| *A. Performance evaluation on the argumentation strength of evidence sentences*<br>*(N = 100, average = 'macro')* |  |  |  |  |
| --- | --- | --- | --- | --- |
| Model | Accuracy | Recall | Precision | F1 (macro) |
| GPT-3.5, few-shot | 0.44 | 0.58 | 0.55 | 0.45 |
| Llama2-7b (576 training set) | **0.84** | **0.68** | **0.91** | **0.70** |
| *B. Performance evaluation on the causal extraction using ChatGLM* |  |  |  |  |
| Test datasets | Relation-entity |  | Entity-relation |  |
|  | Recall | Precision | Recall | Precision |
| GDM (223 sentences, 2202 items) | 85.20% | **86.36%** | 85.20% | 85.97% |
| NLM GoldStandard (142 sentences) | 76.06% | 76.6% | **85.21%** | 85.21% |
| NLM SemRep (factuality, 1635 sentences) | 65.32% | 66.67% | 70.64% | **71.21%** |

*Note:* For each method, the results in bold are the best scores. Bert F1 score = 0.6; NLM gold standard causal triples

## Discussion and Conclusion

The present study presents an LLMs-enabled causal argument mining framework for casual graph construction and evidence integration. Inspired by argument mining theory, we propose a causal framework consisting of five key elements, which are argument cogency, concept, relation stance, claim-context relevance, and conditional information. Based on the framework, we integrated the fine-tuned Llama2-7b (accuracy, 0.84) for evidence cogency and ChatGLM and PubTator for the causal and conclusive triples extraction (recall, 85.21%), exhibiting a great performance in each task. In particular, the fine-tuned Llama2-7b outperforms few-shot GPT-3.5 in argument cogency and the entity-relation technique exceeds the relation-entity one in casual extraction.

The causal framework and LLM-enabled automatic extraction reveal referrable method details in future causal mining tasks. First, by transitioning conclusive semantic triples in the strong persuasive statement into causal relations, we provide a feasible thought and methodology for defining and detecting causality from the perspective of argumentation and text mining. Second, compared to GPT-3.5 turbo with few-shot and GPT-4 with few-shot, the excellent performance of the fine-tuned Llama2-7b model using merely 576 high-quality annotation sentences proves the feasibility and capacity on a moderately difficult task by fine-tuning lightweight models with a small training set. This is of great significance and benefits to make it feasible to be applied to a large scale of texts, which extremely increases the accuracy of performance, saves computational costs, and improves its applicability. Third, by combining PubTator and ChatGLM, the prominent entity-relation extraction approach provides a solution to entity alignment, which can better link to downstream causal graph-built tasks. Though a brilliant understanding of complex texts and relations, LLMs-based extraction techniques are used to encounter the tough entity alignment problem due to its borderless phrase detection in NER tasks. Identifying standard concepts first by PubTator and causal relations later makes it much easier than mapping ChatGLM extracted irregular length concept phrases to standard concepts.

Though promising results are revealed, limitations exist in the study. First, limited LLMs are considered and compared in this work. To test the complexity and feasibility of the framework, we choose state-of-the-art models for the initial trial. Except for trials and baseline comparison, we mainly focus on open-source LLMs for the availability of real applications. Second, as we use the SemMedDB dataset, the contexts are only sentences in the title and abstract of an article. Though abstracts are representative covering all sections from background to conclusion, future work will consider including full-text bodies to further evaluate models and optimize the prompt engineering. Third, considering the inherent feature of the GDM validation set, we only consider one conditional information (the research object) in our evaluation. For diseases with different traits, one may consider designing a different conditional information extraction schema.

Collectively, for a given pair of exposure-outcome (subject-object) of interest, by automatically integrating all conclusive triples of cogent evidence, one can delineate the whole picture of the causal argument structure of the given two concepts. The LLMs-enabled causal extraction approach serves as a crucial foundation for various downstream tasks, such as the evidence integration of exposures and outcomes, intermediating variables mining based on the A-B-C model of literature-based discovery, and the construction and various applications of causal graphs.

## Data Availability

All data produced in the present study are available upon reasonable request to the authors

## Acknowledgments

This study was funded by the National Key R&D Program for Young Scientists (Project number 2022YFF0712000 to JD) and the National Natural Science Foundation of China (Project number 72074006 to JD).

